# Pattern and mechanisms of atrophy progression in individuals with a family history of Alzheimer’s disease: a comparative study

**DOI:** 10.1101/2024.03.01.24303606

**Authors:** Christina Tremblay, Shady Rahayel, Alexandre Pastor-Bernier, Frédéric St-Onge, Andrew Vo, François Rheault, Véronique Daneault, Filip Morys, Natasha Rajah, Sylvia Villeneuve, Alain Dagher, the PREVENT-AD Research Group, Alzheimer’s Disease Neuroimaging Initiative (ADNI)

## Abstract

Alzheimer’s disease (AD) includes a long period of presymptomatic brain changes. Different risk factors are associated with AD development, including having a family history of AD (FHAD). The Braak scheme suggests that tau pathology, in synergy with amyloid-beta (Aβ), spreads along structural connections in AD, eventually leading to atrophy. Studying the pathways in which atrophy spreads early on, as well as the factors underpinning this pathway, is crucial for improving diagnostic accuracy and early interventions. However, the pattern of atrophy progression in people with a FHAD and the biological factors associated with this progression remain unclear. Here we used structural MRI from three databases (ADNI, PREVENT-AD and Montreal Adult Lifespan Study) to map the atrophy progression in FHAD and AD and assess the constraining effects of structural connectivity on atrophy progression. Cross-sectional and longitudinal data up to 4 years were used to perform atrophy progression analysis in FHAD and AD compared to controls. Positron emission tomography (PET) radiotracers were also used to quantify the distribution of tau and Aβ proteins at baseline. We first derived cortical atrophy progression maps using deformation-based morphometry from 153 FHAD, 156 AD, and 116 controls with similar age, education, and sex at baseline. We next examined the spatial relationship between atrophy progression and spatial patterns of tau and Aβ deposition, structural connectivity, and neurotransmitter receptor and transporter distributions. Our results show that there were similar patterns of atrophy progression in FHAD and AD, notably in the cingulate, temporal and parietal cortices, with more widespread and severe atrophy in AD. Both tau and Aβ pathology tended to accumulate in regions that were structurally connected in FHAD and AD. The pattern of atrophy and its progression also aligned with existing structural connectivity in FHAD. In AD, our findings suggest that atrophy progression results from propagating pathology that occurred much earlier, on an intact connectome. Moreover, a relationship was found between the serotonin 5-HT6 receptors spatial distribution and atrophy progression in AD, supporting an important role of these receptors in neurodegeneration. The current study demonstrates that regions showing atrophy progression in FHAD and AD present with specific connectivity and cellular characteristics, uncovering certain of the mechanisms involved in preclinical and clinical neurodegeneration.

## Introduction

In Alzheimer’s disease (AD), family history stands as the second strongest risk determinant, surpassed only by advanced age (Tanzi, 2012). A meta-analysis revealed a 3.5-fold increase in AD susceptibility for individuals with at least one first-degree relative with AD (“family history of AD”, FHAD) (Van Duijn et al., 1991). The heritability of AD is estimated to be between 58% and 79% (Karlsson et al., 2022)) and one of the most well-established genetic risk factors is the presence of one or two e4 alleles of the apolipoprotein E gene (*APOe4*) (M Donix et al., 2012). However, the etiology of AD extends beyond a single genetic locus and *APOe4* interacts with other genetic and environmental factors to influence AD risk as well as brain structure and function (Andrews et al., 2023). The concept of family history as a risk factor may encapsulate both known and cryptic genetic susceptibilities, as well as non-genetic factors that are transmissible across generations (Borenstein et al., 2006; Robinson et al., 2008). Investigating how brain structures are affected in individuals with FHAD could offer insights into the neuronal mechanisms underlying the onset of atrophy in AD or its alleviation.

AD pathology is associated with brain structural alterations and tissue loss across various regions, notably the temporal lobe, frontoparietal and parieto-occipital regions, and the hippocampus (Bakkour et al., 2013; Binette et al., 2020; Du et al., 2001; Jagust et al., 2008). The medial temporal lobe and hippocampus are among the earliest regions to show atrophy, while other cortical regions are affected at later stages (Planche et al., 2022). While hippocampal atrophy has been extensively studied in the context of AD (Barnes et al., 2009; Shi et al., 2009), it lacks specificity as it also occurs in other forms of dementia such as vascular, semantic and Parkinson’s disease dementia (Pini et al., 2016). Therefore, cortical atrophy patterns may offer a more specific marker for tracking AD progression, especially in its preclinical stages (Pini et al., 2016). Impaired white matter integrity in widespread brain regions (decreased in fractional anisotropy and increase mean diffusivity) (Sexton et al., 2011), white matter loss and reduction in fibre density in specific pathways (Mito et al., 2018) and decreased structural connectivity between brain regions (Palesi et al., 2016), have also been observed in AD. On the other hand, brain structural changes in FHAD remain elusive and have been under investigated. In FHAD, whole-brain gray matter atrophy (Honea et al., 2011) and local atrophy in the precuneus and insula (Kate et al., 2016) have been reported, alongside white matter damage (lower fractional anisotropy, higher mean and radial diffusivity) in regions affected in AD, including the cingulum and uncinate fasciculus (Bendlin et al., 2010; Binette et al., 2021). Atrophy in FHAD has been associated with higher tau and amyloid-beta (Aβ) depositions (Binette et al., 2021). However, the progression of atrophy and its modulation by structural connectivity and, underlying tau and Aβ depositions in FHAD remain unclear.

Alterations in neurochemical systems (such as dopamine, norepinephrine, serotonin, acetylcholine, glutamate and histamine) are well known in AD and may contribute to brain atrophy (Reddy, 2017; Xu et al., 2012). Notably, serotonin depletion has been shown to be related to pathology, depression and sleep disturbances in AD (Reddy, 2017). In addition, loss of acetylcholine-producing neurons can lead to cognitive decline and histaminergic dysregulation can contribute to brain inflammation (Reddy, 2017). Also, reduction of dopamine receptors has been associated with cognitive deficit severity in AD (Xu et al., 2012) while norepinephrine reduction has been linked with neurotoxic proinflammatory conditions and reduces Aβ clearance (Chalermpalanupap et al., 2013). Excessive glutamate release can also lead to neurotoxicity and contribute to neural degeneration (Reddy, 2017). However, the specific mechanisms implicated in AD-related brain atrophy progression is still an open question and which neurotransmitter systems interact in FHAD to influence atrophy progression remains to be elucidated.

Therefore, brain atrophy in AD might be influenced by multiple factors, including accumulation of tau and Aβ deposition, reduced structural connectivity, as well as changes in neurotransmitters distribution. Understanding how these factors role shape the trajectory of brain atrophy progression in individuals with FHAD could be key to early detection and intervention. For example, treatments could be developed to reduce tau and Aβ deposition, to block the spread of pathologocial agent throught the brain connectome or enhance specific neurotransmitter functions. The primary aim of this study was to elucidate the features underlying cortical atrophy progression in individuals with a FHAD and compare them to those in individuals with AD. We applied Deformation-Based Morphometry (DBM) to quantify cortical atrophy progression in both groups and advanced tractography algorithms to assess whether structural connectivity exerted a constraining effect on brain atrophy progression, tau, and Aβ pathology in each group. Using PET scans, we next investigated whether the atrophy progression patterns significantly overlapped the spatial distributions of tau and Aβ pathology. Finally, we studied whether brain atrophy progression patterns occurred within regions expressing specific types of neurochemical receptors and transporters based on curated PET maps.

## Materials and methods

We employed T1-weighted MRI to quantify brain atrophy progression, Tau-PET and Aβ-PET via Positron Emission Tomography for mapping Alzheimer-related pathology, and Diffusion-Weighted Imaging (DWI) to quantify cortical structural connectivity in FHAD and AD. Figure 1 provides a summary of the main methodological steps for each of these three modalities.

**Figure 1.**
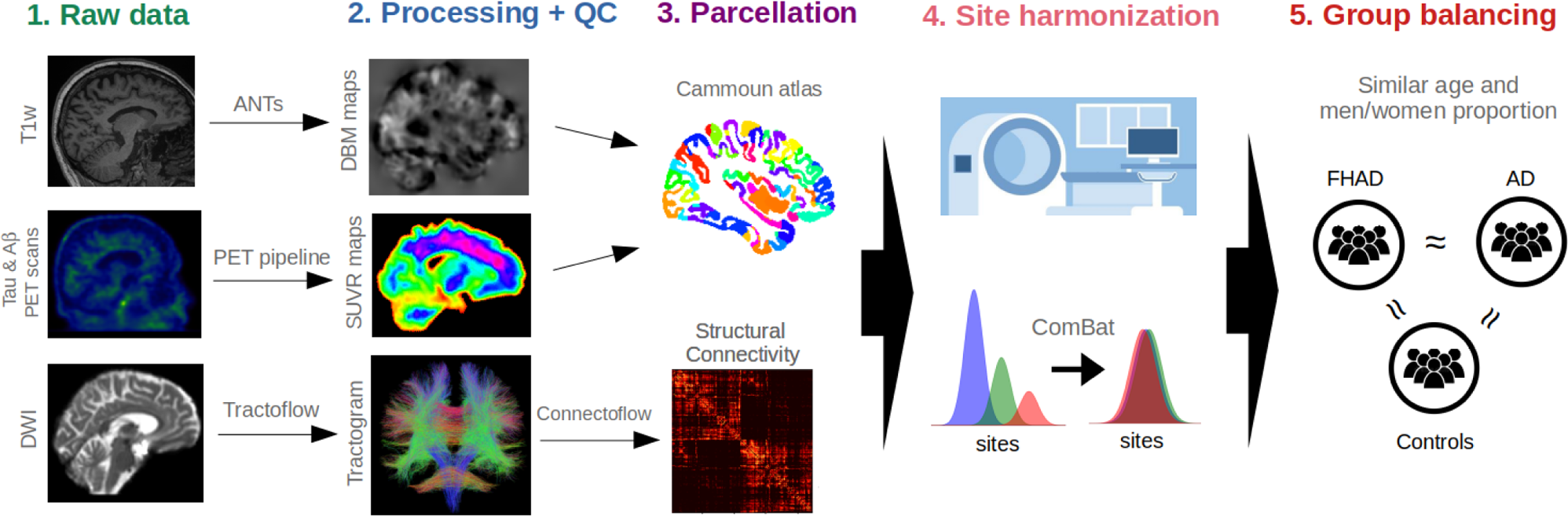
Five main steps of the method for the three modalities used in this study. The data were first acquired from three different databases (step 1). Then, all neuroimaging data were processed using different software specific to each modality, followed by quality control (QC) (step 2). Subsequently, each brain map was parcellated using the Cammoun atlas with 448 cortical regions (step 3), and the site effect was regressed out using the ComBat software (step 4). Lastly, we ensured that each group (participants with a family history of Alzheimer’s disease (FHAD) or with Alzheimer’s disease (AD) and healthy controls (when applicable)) had a similar age and men/women proportion at baseline (step 5).

### 1 Brain atrophy progression

#### 1.1 Datasets

To create cortical atrophy progression maps for individuals with FHAD, AD dementia, and cognitively unimpaired healthy control (HC) without a FHAD, clinical and structural MRI data were downloaded from three databases: the Presymptomatic Evaluation of Experimental or Novel Treatments for Alzheimer’s Disease (PREVENT-AD), internal and open database (Breitner et al., 2016), The Alzheimer’s Disease Neuroimaging Initiative (ADNI) (Jack et al., 2008), and the Montreal Adult Lifespan Study (MALS) (Ankudowich et al., 2016) in August 2021. Adults (> 55Y) with T1-weighted MRI at baseline (in the internal PREVENT-AD, ADNI and MALS databases) and follow-up MRIs over a period of up to four years (in the internal PREVENT-AD and ADNI databases) were included in this study. The number of participants at each time point and each main step of the method are detailed in Table 1 (see Supplementary Table.1 for the number of subjects at each step in each database separately). The acquisition parameters for the PREVENT-AD and MALS datasets (internal databases) were the same. The detailed protocols are described in (Tremblay-Mercier et al., 2021). All MRI images from these two databases were acquired on a Magnetom Tim Trio 3 Tesla (Siemens) scanner at the Douglas Mental Health University Institute (Montreal, Canada). PREVENT-AD is a longitudinal study of cognitively normal older adults who have a parent or at least one sibling diagnosed with AD (FHAD). Participants enrolled in the study were required (1) to be at least 60 years old, or between 55 and 59 if their age was within 15 years of their first-affected relative’s age at the onset of dementia, (2) to not have a history of neurological or psychiatric disorders, and (3) to have normal cognitive functions as indicated by a neuropsychological evaluation. In the MALS database, all participants were healthy adults (age range: 19-76Y) with no history of neurological or psychological illness or family history of AD.

**Table 1.** Number of subjects at each main step of the method for the patients with Alzheimer’s disease (AD) and the individuals with a family history of AD (FHAD)

| Data | Group | Database | Raw data |  | Processing + QC |  | Parcellation | Site harmonization |  | Group balancing |  |
| --- | --- | --- | --- | --- | --- | --- | --- | --- | --- | --- | --- |
| ATROPHY PROGRESSION - DEFORMATION-BASED MORPHOMETRY (DBM) |  |  |  |  |  |  |  |  |  |  |  |
| T1w-MRI | AD | ADNI | Bl: 418 | 1Y: 237 | Bl: 340 | 1Y: 190 | No data removed | No data removed |  | Bl: 156 | 1Y: 96 |
|  |  |  | 2Y: 153 | 3Y:18 | 2Y: 121 | 3Y:17 |  |  |  | 2Y: 55 | 3Y: 5 |
|  | FHAD | ADNI PREVENT-AD | Bl: 320 | 1Y: 216 | Bl: 280 | 1Y: 148 | No data removed | No data removed |  | Bl: 153 | 1Y: 126 |
|  |  |  | 2Y: 179 | 3Y: 65 | 2Y: 155 | 3Y: 53 |  |  |  | 2Y: 110 | 3Y: 46 |
|  |  |  | 4Y: 99 |  | 4Y: 83 |  |  |  |  | 4Y: 65 |  |
|  | HC | ADNI MALS | Bl: 153 | 1Y: 70 | Bl: 131 | 1Y: 55 | No data removed | Bl: 127 | 1Y: 55 | Bl: 116 | 1Y: 47 |
|  |  |  | 2Y: 70 | 3Y: 48 | 2Y: 56 | 3Y: 39 |  | 2Y: 56 | 3Y: 39 | 2Y: 48 | 3Y: 33 |
|  |  |  | 4Y: 27 |  | 4Y: 20 |  |  | 4Y: 20 |  | 4Y: 18 |  |
| POSITRON EMISSION TOPOGRAPHY IMAGING (PET) |  |  |  |  |  |  |  |  |  |  |  |
| Tau-PET | AD | ADNI | Bl: 64 |  | Bl: 62 |  | No data removed | Bl: 58 |  | Bl: 58 |  |
|  | FHAD | ADNI PREVENT-AD | Bl: 166 |  | Bl: 157 |  | No data removed | Bl: 146 |  | Bl: 96 |  |
| Aβ-PET | AD | ADNI | Bl: 167 |  | Bl: 159 |  | No data removed | Bl: 154 |  | Bl: 145 |  |
|  | FHAD | ADNI PREVENT-AD | Bl: 221 |  | Bl: 219 |  | No data removed | Bl: 214 |  | Bl: 165 |  |
| STRUCTURAL CONNECTIVITY - DIFFUSION TENSOR IMAGING (DTI) |  |  |  |  |  |  |  |  |  |  |  |
| DWI-MRI | AD | ADNI | Bl: 116 |  | Bl: 91 |  | Bl: 90 | No data removed |  | Bl: 72 |  |
|  | FHAD | PREVENT-AD | Bl: 304 |  | Bl: 277 |  | Bl: 274 | No data removed |  | Bl: 78 |  |

QC: Quality Control, CR: cortical regions, Bl: Baseline, Y: Year

Participants from the ADNI database (adni.loni.usc.edu) were assigned to one of three groups for our analysis: HC, FHAD, or AD. The ADNI was launched in 2003 as a public-private partnership, led by Principal Investigator Michael W. Weiner, MD. The primary goal of ADNI has been to test whether serial MRI, PET, other biological markers, and clinical and neuropsychological assessment can be combined to measure the progression of mild cognitive impairment (MCI) and early Alzheimer’s disease (AD). For up-to-date information, see www.adni-info.org. Participants with Mini-Mental Status Exam (MMSE) scores from 24 to 30, a normal delayed recall of 1 paragraph from the Logical Memory II subscale of the Wechsler Memory Scale–Revised and a Clinical Dementia Rating (CDR) score of zero were assigned to the HC group if they had no family history of AD, or to the FHAD group if they had a parent or a sibling with AD. ADNI participants with an MMSE score from 20 to 26, impairments on the delayed recall of 1 paragraph from the Logical Memory II subscale of the Wechsler Memory Scale–Revised, a CDR score ≥ 0.5 and who met NINCDS/ADRDA criteria for probable AD (McKhann, 1984) were assigned to the AD group. The inclusion criteria in ADNI were described previously (Petersen et al., 2010). Exclusion criteria in ADNI included any significant neurological disease other than AD, major depression, bipolar disorder, a history of schizophrenia, and a history of alcohol or substance abuse within the past 2 years. T1-weighted MRI from ADNI, acquired on 3 and 1.5 Tesla scanners, were included (detailed protocol described in (Jack et al., 2008)).

For all datasets, participating centers received approval from a local research ethics committee. All the procedures and tests followed these committees’ guidelines. Informed consent was obtained from each participant according to the Declaration of Helsinki before the beginning of each study.

#### 1.2 Processing and Quality Control

DBM was used as a voxel-wise measure of brain atrophy. DBM maps were derived from each participant’s T1-weighted MRI image at each time point by performing non-linear transformations from the participant’s brain to a template (MNI152-2009c) using the Advanced Normalization Tools (ANTs) Longitudinal Cortical Thickness Pipeline (Tustison et al., 2014, 2018). DBM maps were generated by concatenating the non-linear warps that mapped the T1-weighted images from each time point to the corresponding subject-specific template, and then by mapping the subject-specific template to the MNI152-2009c template. The derivative of these deformation maps was used to estimate local tissue volume changes, which were quantified as the determinant of the Jacobian matrix of displacement. The maps were smoothed with a 2 mm Gaussian kernel to decrease spatial noise. Finally, the natural logarithm of the Jacobian determinant was calculated. Relative to the MNI template, a value of zero indicates no volume difference, negative values indicate tissue expansion and positive values indicate tissue loss (atrophy). Quality control was done by visual inspection of the resultant DBM maps. In total, 158 subjects with AD (19%), 40 FHAD subjects (18%) and 67 HC (18%) were excluded mostly due to segmentation issues (see Table 1 for the remaining number of subjects at each time point).

#### 1.3. Parcellation, site harmonization, and group balancing

DBM maps were parcellated using the Cammoun atlas consisting of 448 equal-size cortical regions (Cammoun et al., 2012). To account for scanning site variability, we used a variational Bayes harmonization method developed for longitudinal brain imaging data. Longitudinal ComBat (LongComBat) was applied region-wise to harmonize the DBM data and regress out site-specific effects (age, sex and group were used as covariates). LongComBat has been validated and shown to be effective in reducing site-specific effects with longitudinal data (Beer et al., 2020). The baseline maps from four subjects with AD were excluded because they were the only subjects for their site. In addition, participants were selected to ensure that the three groups were similar in terms of age (mean FHAD: 72.6Y, range: 66 - 87.8Y, mean AD: 72.8Y, range: 55.2 - 83.7Y and HC: 72.6Y, range: 60.0 - 83.4Y; p-value=0.94), education (mean FHAD: 16.1Y, range: 9 - 20Y, mean AD: 15.6Y, range: 13 - 20Y, and HC: 16.1Y, range: 7 - 20Y; p-value=0.15) and sex (FHAD vs. HC: χ2 = 3.67, p-value= 0.06; AD vs. HC: χ2 = 3.67, p-value= 0.06) at baseline. Individuals with a FHAD who were under 66 years old and, HC and AD subjects over 84 years of age were removed. Ultimately, this study included data from 153 subjects with FHAD (85 women), 156 subjects with AD (80 women), and 116 HC (73 women) at baseline, with follow-ups for up to four years.

#### 1.4 Participants

Table 2 describes the clinical characteristics of the participants with AD, FHAD, and HC at baseline included in this study. Two-tailed F-tests with post-hoc comparisons and Bonferroni-Holm correction were used to compare the groups. Compared to HC, the Montreal Cognitive Assessment (MoCA) score (Nasreddine et al., 2005), corrected for education, was significantly lower in AD (mean AD: 16.3 vs. HC: 27.1; *p-value*<.0001) as well as the MMSE score (mean AD: 23.1 vs. HC: 29.2; *p-value*<.0001). Slightly higher depressive symptoms as assessed through the GDS score (Marc et al., 2008) were found in AD (mean AD: 1.7 vs. HC: 0.8; *p-value*<.0001). A higher proportion of AD subjects (70%) carried either one or two APOe4 alleles compared to HC (26%), while a slightly higher proportion of FHAD subjects carried two APOe4 alleles (FHAD: 8.6% vs. HC: 0.9%).

**Table 2.** Descriptive statistics for the controls and participants with a family history of Alzheimer’s disease (FHAD) and Alzheimer’s disease (AD) included in this study.

| Characteristics | Baseline Control | Baseline FHAD | Baseline AD | F-test | FHAD vs Control | AD vs Control |
| --- | --- | --- | --- | --- | --- | --- |
|  | mean (SD) | mean (SD) | mean (SD) | P-Value Adjusted* | p-value | P-value |
| Number of subjects (Nb) | 116 | 153 | 156 | NA | NA | NA |
| Gender (men/women)** | 43/73 | 68/85 | 76/80 | NA | 0.22 | 0.06 |
| Age (years) | 72.6 (6.4) | 72.6 (5.0) | 72.8 (6.7) | 0.94 | NA | NA |
| Education (years) | 15.6 (2.5) | 16.1 (2.6) | 16.1 (2.2) | 0.30 | NA | NA |
| MMSE*** | 29.17 (0.98) | 29.08 (1.05) | 23.11(2.22) | **2x10<sup>-120</sup>** | 0.68 | **1.7x10<sup>-103</sup>** |
| MoCA | 27.09 (2.86) | 26.12 (2.44) | 16.33 (4.72) | **2x10<sup>-64</sup>** | 0.06 | **1x10<sup>-55</sup>** |
| Depression (GDS)*** | 0.78 (1.19) | 0.75 (1.17) | 1.70 (1.49) | **1x10<sup>-8</sup>** | 0.87 | **3x10<sup>-6</sup>** |
| APOE4 (% of subjects with the gene in 1 allele)** | 25% | 27% | 49% | NA | 0.72 | **0.0001** |
| APOE4 (% of subjects with the gene in 2 alleles)** | 0.9% | 8.6% | 21% | NA | **0.005** | **< 0.00001** |

\*P-adjusted: Bonferroni-Holm correction
\*\* $\chi^2$ test \*\*\*Data from ADNI cohort: data available for 59% (Control) and 70% (FHAD) of the subjects
NA: Not applicable; MoCA: Montreal Cognitive Assessment; SD:Standard Deviation; GDS: Geriatric Depression Scale

#### 1.5 Statistical analysis

The pattern of brain atrophy progression in FHAD and AD was investigated using linear mixed models with random intercept and slope. The atrophy progression (group*age interaction) was compared region-wise between the three groups (FHAD, AD and HC). Group, age, sex, education, body mass index (BMI), *APOe4* status, and *APOe4*\*age interaction were used as covariates. Post-hoc tests were also computed between each group. This analysis was performed using MATLAB (R2020b). Secondary analyses were done to further assess the effect of sex by adding a group*sex*age interaction, as well as group*sex and sex*age interactions, in the models. False Discovery Rate (FDR) corrections were applied to control for multiple comparisons (Benjamini & Yekutieli, 2005). In all subsequent analyses, the regional b-values (beta) of the group*age interaction (FHAD vs. HC and AD vs. HC) were used as an estimate of region-wise atrophy progression in FHAD and AD. A positive b-value indicates more brain atrophy progression compared to HC, while a negative b-value indicates less atrophy progression. In addition, W-Score maps were generated region-wise for brain atrophy at baseline to account for the effects of age and sex (La Joie et al., 2012). A higher W-Score indicates greater atrophy at baseline compared to HC. Pearson’s correlations between FHAD and AD atrophy progression, and atrophy at baseline, were also computed. Their significance was tested against spatially auto-correlated null models using the software BrainSmash (1000 spins, two-tailed) (Burt et al., 2020; Markello & Misic, 2021).

#### 1.6 Brain atrophy progression in the resting-state networks

To explore the relationship between brain atrophy and cognitive functions, the mean atrophy progression was calculated for each of the seven cortical resting-state networks, as defined by (Yeo et al., 2011). The mean atrophy progression in each network was compared between the three groups (FHAD, AD, and HC) with linear mixed models (random intercept and slope), along with post-hoc tests. Group, age, sex, education, BMI, *APOe4* status and *APOe4*\*age interaction were used as covariates. Partial Spearman’s correlations, adjusting for age at baseline, BMI and *APOe4* status, were also performed to assess the associations between global cognitive measures (education-corrected MoCA scores) and baseline atrophy (as indicated by W-Scores regressing out normal aging and sex effect) across each of the seven resting-state networks (*p-value_FDR_*<.05, two-tailed).

### 2 Positron Emission Topography Imaging (PET)

#### 2.1 Datasets

To create the tau-PET and Aβ-PET maps at baseline, PET scans from the PREVENT-AD (internal database) and ADNI were acquired for subjects with FHAD (tau:166 scans; Aβ:221 scans) and AD (tau:64 scans; Aβ:167 scans) (Table. 1). Only one scan of each tracer per subject (baseline) was included in this study for both tau and Aβ, since there were no follow-up scans in the PREVENT-AD study, and the number of follow-up scans was low and inconsistent between FHAD and AD in ADNI. We obtained the unprocessed PET data for each acquisition in ADNI.

For the subjects with FHAD from the PREVENT-AD database, the radiotracers [18F]AV-1451 (flortaucipir) and [18F]NAV4694 were used to quantify the distribution of tau and Aβ in the brain (tau: 120 scans; Aβ: 122 scans). PET scans were performed at the McConnell Brain Imaging Centre at the MNI (Montreal, Canada) using a brain-dedicated PET Siemens/CT high-resolution research tomograph. Tau scans were performed 80 to 100 min after radiotracer injection (9.9 ± 1.0 mCi) and Aβ scans were performed 40 to 70 min after injection (6.6 ± 0.4 mCi). T1-weighted MRI scans were acquired up to one year before the PET scans (mean interval: 8.9 ± 4.8 months) on a 3T Siemens Trio scanner at the Brain Imaging Centre of the Douglas Mental Health University Institute (Montreal, Canada). The following parameters were used: TR: 2300 ms, TE: 2.98 ms, FA: 9°; matrix size: 256 x 256; voxel size: 1 mm^3^; 160-170 slices.

Baseline PET data from the ADNI database were downloaded in January 2023. The radiotracers [18F]AV-1451 (flortaucipir) and [18F]AV-45 (florbetapir) were used to assess tau and Aβ distribution in subjects with FHAD and AD. This dataset included 46 tau and 99 Aβ scans for FHAD, and 64 tau and 167 Aβ scans for AD (Table.1). T1-weighted MRI images, acquired up to one year prior to the PET scans, were also downloaded. Acquisition parameters and detailed protocols for the T1-weighted MRI are described in (Jack et al., 2008). For flortaucipir, six five-minute frames were acquired starting 75 minutes following radiotracer injection (10.0 ± 1.0 mCi). For florbetapir, four five-minute frames were acquired starting at 50 minutes following radiotracer injection (10.0 ± 1.0 mCi). PET scanners slightly differed by site; more information on PET image acquisition in the ADNI database is described in detail at http://adni-info.org.

#### 2.2 Processing and Quality Control

PET scans from the PREVENT-AD and ADNI databases were then processed with the same standard pipeline (github.com/villeneuvelab/vlpp) and quality control. Briefly, for each participant, the PET image frames were realigned, averaged, and registered to the corresponding T1-weighted MRI processed using FreeSurfer v.6.0. Registered PET images were then masked to exclude CSF signal and finally smoothed (Sperling et al., 2011). For the smoothing, a 6 mm Gaussian kernel was used for PREVENT-AD scans, while a 8 mm kernel was applied for ADNI scans to match the approximate resolution of the lowest resolution scanners used (Jagust et al., 2015). Standardized uptake value ratios (SUVRs) using the inferior cerebellum gray matter for tau scans (Baker et al., 2017) and cerebellum gray matter as the reference region for Aβ scans (Villeneuve et al., 2015) were computed voxel-wise. PET and T1w images were coregistered and mapped into the MNI space. Quality control was performed to remove PET and T1-weighted images with artifacts, segmentation or reconstruction issues, and misalignment between PET and T1w images. No participants were excluded from the PREVENT-AD cohort. From the ADNI database, 9 tau (5%) and 2 Aβ (1%) scans from FHAD subjects, along with 2 tau (3%) and 8 Aβ (5%) scans from AD subjects, were excluded.

#### 2.3 Parcellation, site harmonization, and group balancing

The Cammoun atlas (Cammoun et al., 2012) was used to parcellate the MRI into 448 equal-sized cortical regions. These regions were then used to extract SUVRs from the PET maps. Then, ComBat was used to regress out inter-site variability from the PET images with age, sex and group as covariates (Johnson et al., 2007). Data from sites with only one subject were excluded: FHAD: tau (N=11) and Aβ (N=5); AD: tau (N=4) and Aβ (N=5). To ensure that the average age of the FHAD group was similar to the average age in the brain atrophy progression analysis, younger FHAD subjects (<66Y) were excluded (tau-PET: t-value =.43, *p-value*=.67 and Aβ-PET: t-value =-0.87, *p-value*=.38). Older AD subjects (>89Y) were also excluded (tau-PET: t-value =-1.18, *p-value*=.24 and Aβ-PET: t-value =-1.41, *p-value*=.16). After site harmonization and group balancing, the spatial distribution of Aβ-PET in FHAD was highly correlated between participants from PREVENT-AD and ADNI despite the two databases using different radiotracers (r = 0.84, *p-value_spin_* = 0.001). In total, tau imaging from 96 subjects with FHAD and 58 patients with AD, and Aβ imaging from 165 subjects with FHAD and 145 patients with AD were used to create the PET maps.

### 3 Structural connectivity

#### 3.1 Datasets

To develop group-specific structural connectivity matrices for the FHAD and AD groups, we used diffusion MRI data from the PREVENT-AD (N=304) and ADNI (N=116) databases, respectively (Table.1). For the subjects with FHAD, the baseline diffusion-weighted MRI (DWI) data were downloaded from the open PREVENT-AD database in February 2023. The DWI consisted of one b_0_ image and 64 diffusion-weighted volumes acquired with a b-value of 1000 s/mm^2^ for all subjects. The PREVENT-AD sequence parameters were as follows: Manufacturer = SIEMENS, repetition time (TR) = 9300 ms, echo time (TE) = 92 ms, and voxel size = 2 mm. The DWI data for the AD subjects at baseline were downloaded from the ADNI database in February 2023. All axial DWI data were acquired with an echo-planar imaging sequence. The scan parameters were as follows: Manufacturer = GE MEDICAL SYSTEMS (N=69), Philips Medical Systems (N=7), SIEMENS (N=40); b-value = 583 to 1225 s/mm²; gradient directions = 30 (N=8), 32 (N=6), 41 (N=49), 48 (N=14), 54 (N=22), 126 (N=10); voxel size = 0.91 × 0.91 mm² (N=20), 1.37 × 1.37 mm² (N=49), 2 × 2 mm² (N=44), 2.7 × 2.7 mm² (N=3); TR = 3400 to 16700 ms; TE = 55 to 105 ms; slice thickness = 2.0 mm (N=67) and 2.7 mm (N=49).

#### 3.2 Processing and Quality Control

Tractography and connectomic pipelines were applied to the DWI data to create a binary group connectivity matrix for both the FHAD group (PREVENT-AD) and the AD group (ADNI). The TractoFlow Atlas Based Segmentation pipeline (TractoFlow-ABS: github.com/scilus/tractoflow), with the connectomics profile, was used to create tractography from raw DWI and T1 re-sampled from FreeSurfer v.6.0. (Theaud et al., 2020b, 2020a). The DWI processing included denoising, topup corrections to reduce the brain deformation induced by the magnetic field susceptibility artifacts, eddy-currents correction, N4 Bias Correction followed by the computation of DWI and fiber orientation distribution function (fODF) metrics (Theaud et al., 2020b). For TractoFlow-ABS, the following parameters were selected for diffusion image processing: DTI shells: 0 500 1000 2000; fODF shells: 0 500 1000 2000; FRF value: 10, 3, 3; algorithm: local probabilistic tracking; local seeding mask type: WM/GM interface; number of seeds per voxel: 20; spherical harmonic (SH) order: 6 (<32 gradient directions) and 8 (>=32 directions). A higher SH order allows for more complex diffusion patterns to be represented in the context of a larger number of gradient directions. These parameters allowed to obtaining valid estimations for both databases and all the gradient direction sequences (Schilling et al., 2017). Using dMRIQCpy, we next produced quality control files for each key step (raw data, intermediate DWI and T1 preprocessing, metrics from DWI and tractogram) to remove DWI data and T1w-MRI showing artifacts (Theaud & Descoteaux, 2022). Five subjects with FHAD failed during the Tractoflow-ABS processing, while 22 DWI images (7%) from the FHAD group (PREVENT-AD) and 25 DWI images (22%) from the AD group (ADNI) were removed after quality control.

#### 3.3 Parcellation, site harmonization, and group balancing

We next used Connectoflow v.1.1.0 (github.com/scilus/connectoflow) to build the structural connectome with the Cammoun atlas (448 cortical regions) for both the FHAD and AD groups (Di Tommaso et al., 2017; Kurtzer et al., 2017; Rheault et al., 2021). The tractograms generated by Tractoflow-ABS were employed in the Connectoflow pipeline. The Convex Optimization Modeling for Microstructure Informed Tractography (COMMIT2) was used to assign to each streamline a weight, which was used for removing false positive brain connections. To achieve this, COMMIT2 reconstructs the connectome, providing the optimal explanation for the diffusion-weighted signal, while also reducing the number of invalid streamlines by minimizing the number of bundles. This filtering method has been shown to highly improve the accuracy of the resulting structural connectomes (Schiavi, Petracca, et al., 2020). A connectivity matrix was obtained for 274 subjects (3 subjects failed Connectoflow) with FHAD and 90 subjects with AD (1 subject failed Connectoflow). Since the DWI data from the AD subjects were obtained from different scanner sites, ComBat was used to remove inter-site variability with age and sex as covariates (Fortin et al., 2017; Johnson et al., 2007). Then, younger FHAD (<66Y) and older AD (>84Y) subjects were removed to ensure that subjects used to build the group connectivity matrix had a similar age range to the FHAD and AD subjects in the brain atrophy progression analysis (FHAD: 66 - 88Y; AD: 55 - 84Y). Finally, 78 FHAD (mean: 70Y, SD: 4Y) and 72 AD subjects (mean: 73Y, SD: 7Y) were included to generate the two binary group-average structural connectivity matrices. For each connectivity matrix, connections were retained if they appeared in at least τAvg subjects, where τAvg is the consensus threshold that results in a binary density equal to that of the typical subject (Rubinov & Sporns, 2010). More specifically, the total COMMIT2 weights were sorted in descending order, and the highest weighted connections were selected based on the average number of connections per subject per hemisphere. The inter- and intra-hemispheric connections were calculated separately.

#### 3.4 Network spread analysis

According to the network spread hypothesis, pathology spreads along existing connections between brain regions (Agosta et al., 2015). We tested this hypothesis in both FHAD and AD using the built group-average structural connectivity matrices and an independent matrix from 70 young healthy adults (age = 29 ± 9Y, 43 men) available at https://doi.org/10.5281/zenodo.2872624 (Mišić et al., 2015). A similar approach was previously reported to investigate atrophy spreading through structurally connected regions in isolated REM sleep behavior disorder (iRBD) (Rahayel et al., 2022), Parkinson’s disease (Tremblay et al., 2021; Vo et al., 2023), and schizophrenia (Shafiei et al., 2019). However, all these studies used a general connectivity matrix from young healthy adults (Mišić et al., 2015) and did not include a structural connectivity matrix specific to the pathology being investigated. In this study, we used the connectivity matrix from young healthy adults and the built group-average connectivity matrices from AD and FHAD to compare the results with an intact connectome (removing the possible effect of normal aging and pathology). For the network analysis, we first computed Pearson’s correlations between the brain atrophy progression observed in each of the 448 cortical regions of the Cammoun atlas and the average atrophy progression of their structurally connected neighborhood. Then, the significance of the correlations was tested against a null model preserving the spatial auto-correlation between regions using the BrainSMASH software (1000 spins) (Burt et al., 2020). The same analysis was also computed using the tau-PET and Aβ-PET measures to investigate the protein spreading hypothesis (Vogel et al., 2020). In a supplementary analysis, Pearson’s correlations were also computed with the non-structurally connected neighbors in both FHAD and AD.

### 4 Neurotransmitter receptors and transporters analysis

We next examined if the regional brain atrophy progression in FHAD and AD, as well as atrophy at baseline in FHAD, AD, and HC, related to the spatial distribution of seven different neurotransmitter receptors or transporters potentially implicated in AD. Spatial deformation relative to the MNI152-2009c template was used as baseline atrophy measure (DBM maps at baseline). The receptor and transporter distributions include dopamine (D2), norepinephrine (NET1), serotonin (5-HT1B and 5-HT6), acetylcholine (VAchT), glutamate (mGluR5), and histamine (H3) (Reddy, 2017; Xu et al., 2012). All the distributions were derived from a meta-analysis of PET studies (Hansen et al., 2022). Neurotransmitter receptors or transporters associated with PET tracers with more than one reference (dopamine (D2), glutamate (mGluR5), and acetylcholine (VAchT)) were z-scored, and a weighted average was calculated to account for the varying number of subjects in each study. Pearson’s correlations were tested against null models preserving the spatial auto-correlation of the regions using BrainSMASH (1000 permutations, two-tailed, with FDR correction: *p-value*<.05) (Burt et al., 2020; Markello & Misic, 2021).

### 5 Data availability

T1w-MRI and PET data used in this article were from the internal PREVENT-AD database (release 6.0) and ADNI dataset (available at https://adni.loni.usc.edu/data-samples/access-data/). The DWI data are available online at https://openpreventad.loris.ca (PREVENT-AD) and at https://adni.loni.usc.edu/data-samples/access-data/ (ADNI). All other datasets and software used are available from the sources cited in the Methods. The brain atrophy progression and PET maps, as well as the structural connectivity matrices for the FHAD and AD groups, are available upon reasonable request to the authors.

## Results

### Brain atrophy progression

The DBM maps were used to reproduce the brain atrophy at baseline (Fig.2A) and compare the progression of atrophy over up to four years in 448 cortical regions between the three groups (FHAD, AD, and HC) (Fig.2B, Fig.2C). This analysis showed a significant difference in atrophy progression between groups (group*age interaction) in 138 cortical regions (*p-value_FDR_* < .05). In AD, we found that atrophy progressed significantly with age (+β) in 24 regions part of the cingulate cortex and occipital, temporal, and parietal cortices, compared with age-expected effects in HC. Ten of these regions also showed significant atrophy progression compared with age-expected effects in the FHAD group: including parts of the right occipital (cuneus and lingual gyrus) and frontal cortices (precentral gyrus); the left temporal (transverse temporal gyrus) and parietal cortices (postcentral gyrus and superior parietal lobule); and the bilateral cingulate cortex (caudal anterior and posterior cingulate gyrus). Several regions (n=102) also showed less atrophy progression (−β) in AD than HC, mostly in the temporal and frontal cortices. Twelve of these regions also showed less atrophy progression in FHAD than HC: including parts of the right parietal cortex (precuneus); left insular, temporal (fusiform gyrus (n=2)), frontal (pars triangularis, lateral orbitofrontal, rostral middle frontal) and occipital cortices (lateral occipital gyrus); and bilateral frontal (superior frontal gyrus) and limbic cortices (parahippocampal gyrus). These results indicate a slower atrophy progression at older age within specific regions in AD and FHAD compared to HC. The effect of sex on atrophy progression was also investigated, but no significant effect was found for the group*sex*age and group*sex interactions (*p-value_FDR_* >.07), nor for the sex*age interaction (*p-value_FDR_* >.13). Suggesting a ceiling effect of atrophy in AD, more atrophy at baseline (W-Score) was negatively associated with atrophy progression in this disease (r=-0.40, *p-value*_spin_=.001), but not in FHAD (r=-0.008, *p-value*_spin_=.87).

**Figure 2.**
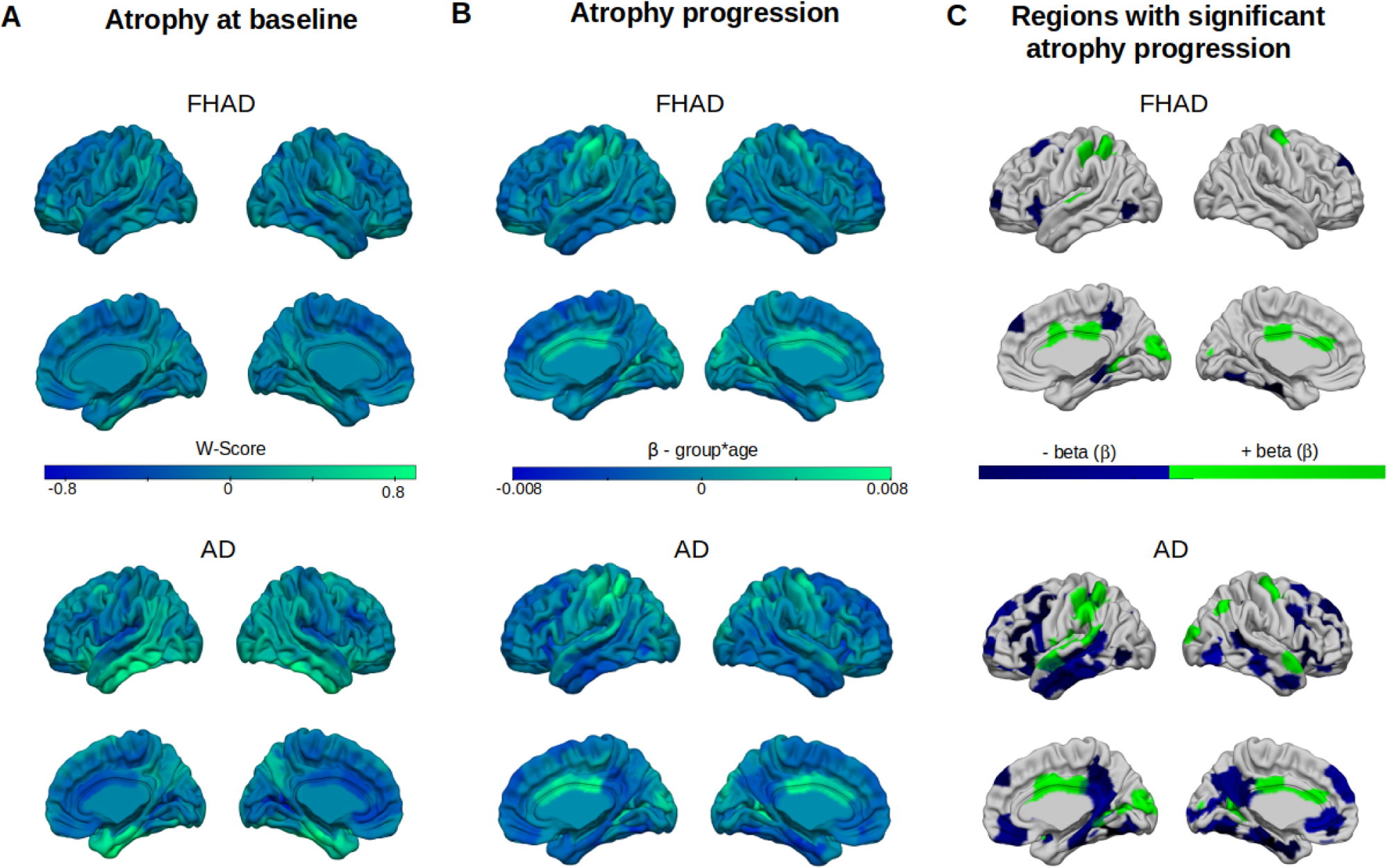
Brain atrophy progression in individuals with a family history of Alzheimer’s disease (FHAD) and patients with Alzheimer’s disease (AD) (**A**) Baseline atrophy (W-scores with age and sex effects in normal aging regressed out) in FHAD and AD. (**B**) Positive (green) and negative (blue) β-values associated with higher and lower atrophy progression in FHAD and AD compared with healthy controls (HC). Regions with more baseline atrophy overlap with regions with less atrophy progression. (C) Cortical regions showing significant atrophy progression (negative atrophy progression in blue, positive atrophy progression in green) in FHAD and AD after FDR corrections.

When comparing AD and FHAD, a significant Pearson’s correlation between AD and FHAD atrophy at baseline (W-Score) was observed (r=0.33, *p-value*_spin_=.001). A significant correlation between groups was also noted for the atrophy progression (r=0.75, *p-value*_spin_=.001). These correlations suggest a similar spatial pattern of atrophy and atrophy progression in both groups (Zou’s CI: −0.51 to −0.33). However, post-hoc comparisons between FHAD and AD revealed that 12 regions, including parts of the bilateral temporal (superior temporal regions) and parietal cortices (supramarginal gyrus), the left occipital (lingual gyrus) and parietal cortices (inferior and superior parietal regions), as well as the right cingulate cortex (postcentral and caudal anterior cingulate gyrus), exhibited significantly greater atrophy progression (+β-value) in AD. Conversely, 80 regions, predominantly located in the temporal, frontal, and parietal cortices, demonstrated significantly less atrophy progression with age (−β-value) in AD (see Supplementary Fig.1 for details). In sum, these results demonstrate that FHAD subjects show brain tissue loss in regions also affected in AD with atrophy being more severe and widespread in AD.

### Brain atrophy progression in the resting-state networks

To describe the association between brain atrophy progression and resting-state functional networks, we quantified atrophy progression and baseline atrophy in the seven resting-state networks as defined by (Yeo et al., 2011). Within these networks, the average atrophy progression was compared between the groups (group*age interaction including the FHAD, AD and HC groups) (Fig.3A). There was a significant interaction for the default mode (*p-value_FDR_*=.00002), limbic (*p-value_FDR_*=.002), somatomotor (*p-value*_FDR_=.008) and ventral attention (*p-value*_FDR_=.03) networks. Post-hoc comparisons showed no significant interaction for FHAD compared to HC. However, the interaction was significant for AD compared to HC in the default mode (β=-.001, *p-value*_FDR_=.00009), limbic (β=-.001, *p-value*_FDR_=.006), and somatomotor (β=.001, *p-value*_FDR_=.01) networks. Only a trend toward significance was observed for the ventral attention network after FDR correction (*p-value*_FDR_=.08). The atrophy progression was not significant in any of the resting-state networks in FHAD compared to HC. However, the results in AD seem to indicate that the atrophy progression had a slower progression at older age or reached a ceiling in the default mode and limbic networks (−β-value), but it is still progressing faster (+β-value) in the somatomotor network.

**Figure 3.**
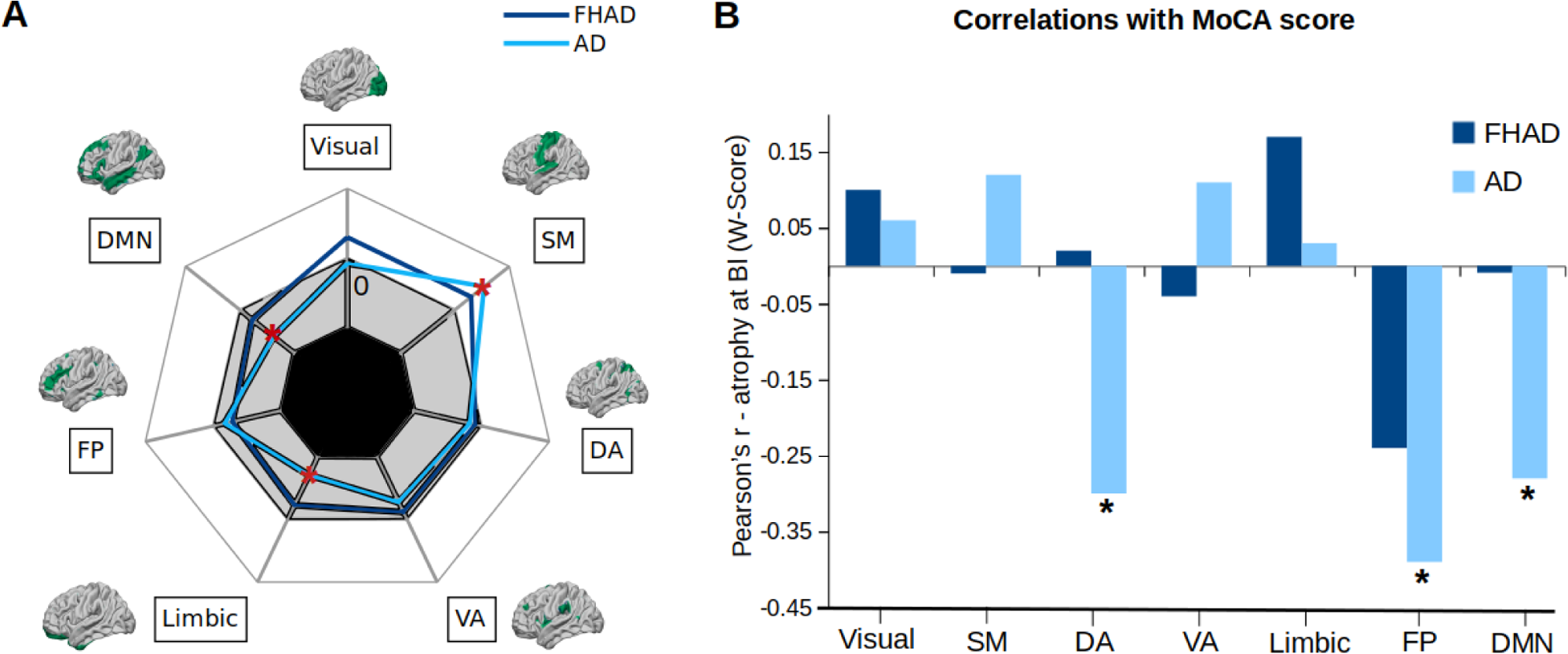
Brain atrophy progression in the seven resting-state Yeo networks in individuals with a family history of Alzheimer’s disease (FHAD) and patients with Alzheimer’s disease (AD) (**A**) Positive and negative β-values associated with higher (white area: b-values range=[0 - 0.002]) and lower (gray area: b-values range=[−0.002 - 0]) atrophy progression in each of the Yeo networks in FHAD and AD. In AD, both the limbic and default mode network (DMN) demonstrated significantly lower atrophy progression compared to healthy controls (HC). Conversely, the somatosensory network showed an increased rate of atrophy progression (\**p-value_FDR_*<.05). (**B**) Pearson’s correlations between average baseline atrophy (W-Score) in each of the Yeo networks and the MoCA score, which evaluates general cognitive abilities, in FHAD and AD. AD participants with higher baseline atrophy in the dorsal attention (DA), frontoparietal (FP), and DMN networks had lower MoCA scores, indicating more cognitive deficits. SM: somatomotor network, DA: dorsal attention network, VA: ventral attention network; FP: frontoparietal network; DMN: default mode network

Moreover, the average atrophy at baseline (W-Score) showed a significant correlation with cognitive abilities (MoCA corrected for education) in the dorsal attention (r=-0.30, *p-value*_FDR_=.02), frontoparietal (r=-0.39, *p-value*_FDR_=.002) and default mode (r=-0.28, *p-value*_FDR_=.02) networks in AD (Fig.3B). No significant correlation was observed in the FHAD group. These results indicate that AD patients with more atrophy in the dorsal attention, frontoparietal or default mode network also have more global cognitive impairment (lower MoCA scores).

### Association with tau-PET and Aβ-PET distribution

Linear regression models were used to compare tau and Aβ concentration between the FHAD and AD groups. These models included age at baseline, sex, education, BMI and *APOe4* status as covariates. After FDR corrections, greater concentrations were found in AD compared with FHAD in 358 cortical regions for tau-PET and 335 regions for Aβ-PET (Supplementary Fig.2). However, both tau and Aβ distribution in FHAD showed significant spatial overlap with tau (r=0.91, *p-value*_spin_=.001) and Aβ (r=0.94, *p-value*_spin_=.001) distribution in AD. This suggests that the patterns of regional distributions are similar in the two groups (Fig.4 upper panel), but that the concentrations of tau-PET and Aβ-PET are higher in AD than FHAD in most cortical regions.

**Figure 4.**
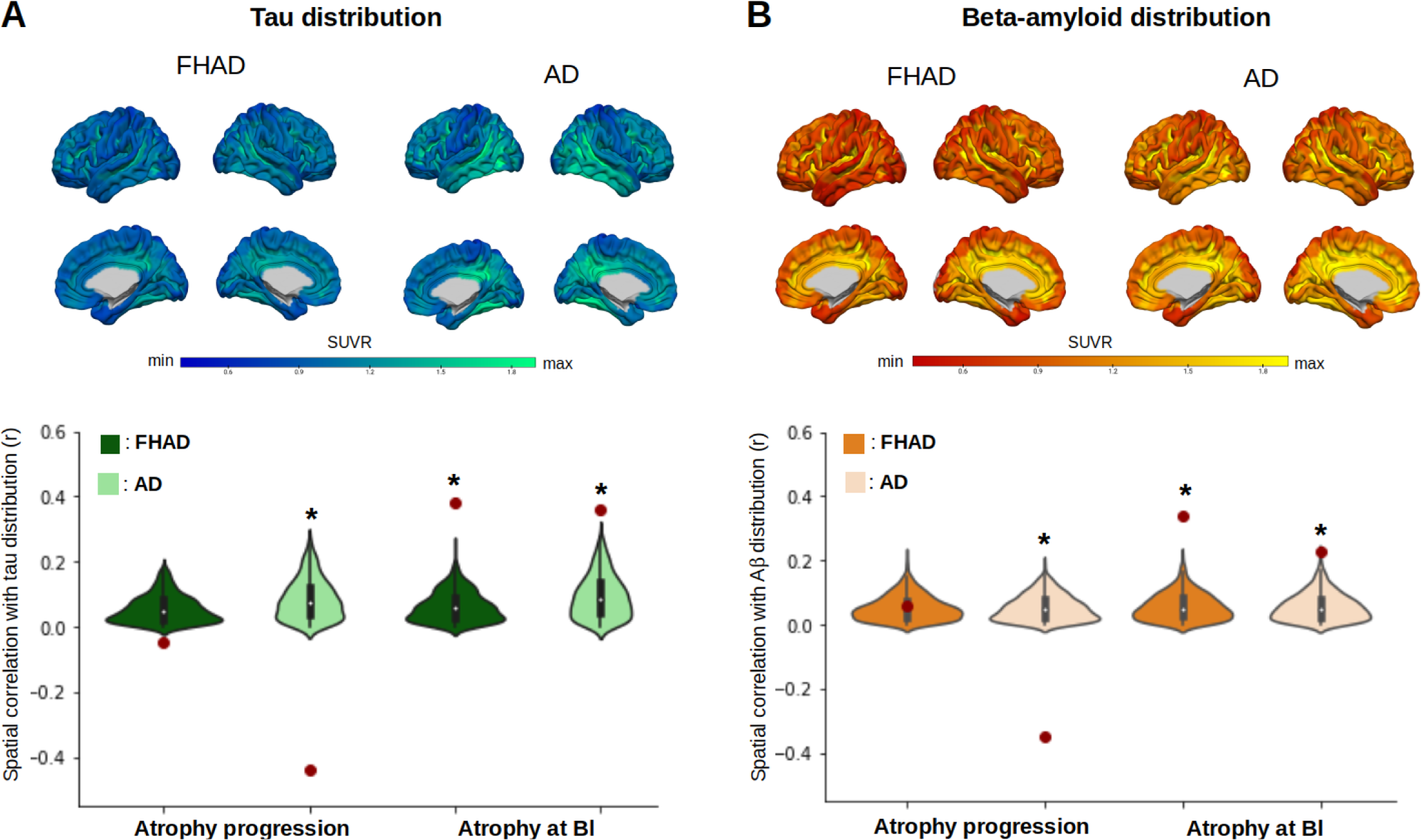
Relationships between brain atrophy, tau and Aβ distribution in individuals with a family history of Alzheimer’s disease (FHAD) and Alzheimer’s disease (AD). The patterns of tau (**A**) and Aβ (**B**) distribution at baseline were significantly and negatively correlated with the atrophy progression only in AD (\**p-value_spin_*<.05). Significant and positive correlations were also found between baseline atrophy and both tau and Aβ distribution in FHAD and AD. All correlations (Pearson’s r) were compared with null coefficient distributions using a model that preserves the spatial auto-correlation between regions.

The relationships of baseline brain atrophy and atrophy progression with baseline tau and Aβ distribution patterns were explored for both FHAD and AD. This analysis was conducted using Pearson’s correlations against spatial null models (Fig.4 lower panel). In FHAD, significant correlations were found between spatial patterns of atrophy at baseline and both tau-PET (r=0.38, *p-value*_spin_=.001) and Aβ-PET (r=0.34, *p-value*_spin_=.001). Similar correlations were obtained when using only FHAD subjects who had both PET and MRI data available at baseline (tau-PET: r=0.40, *p-value*_spin_=.006; Aβ-PET: r=0.39, *p-value*_spin_=.001) (Supplementary Fig.3). However, no significant correlation was observed between the spatial patterns of atrophy progression and tau-PET (r=-0.05, *p-value*_spin_=.26) or Aβ-PET (r=0.06, *p-value*_spin_=.18) distribution in FHAD. In AD, positive and significant correlations were observed between atrophy at baseline and both tau-PET (r=0.36, *p-value*_spin_=.001) and Aβ-PET (r=0.23, *p-value*_spin_=.001). Comparable associations were observed when analyzing only participants with AD who had both PET and MRI data available at baseline (tau-PET: r=0.28, *p-value*_spin_=.001; Aβ-PET: r=0.26, *p-value*_spin_=.002) (Supplementary Fig.3). Moreover, lower atrophy progression with age was related with more tau-PET (r=-0.44, *p-value*_spin_=.001) and Aβ-PET (r=-0.35, *p-value*_spin_=.001) pathology. In sum, both tau and Aβ distribution are related to the atrophy pattern in FHAD and AD, but not to the subsequent atrophy progression in FHAD.

### Association with structural connectivity

We next investigated whether the distribution of cortical atrophy and atrophy progression as well as tau or Aβ spatial distributions could be explained by a process propagating via brain connections (Fig.5). If so, a region’s atrophy, atrophy progression, or tau or Aβ concentration should correlate with the same measure in its structurally connected neighbors. Since structural connections will degrade as the disease runs its course (Agosta et al., 2015), along with an independent structural connectivity matrix from healthy adults, we built structural connectivity matrices from the subjects with FHAD and AD to investigate brain atrophy, atrophy progression, and pathology distributions following the breakdown of connectivity associated with the disease. This allowed us to compare the role of connectivity prior to appearance of pathology (using an intact connectome) and at different stages of the neuropathological process.

**Figure 5.**
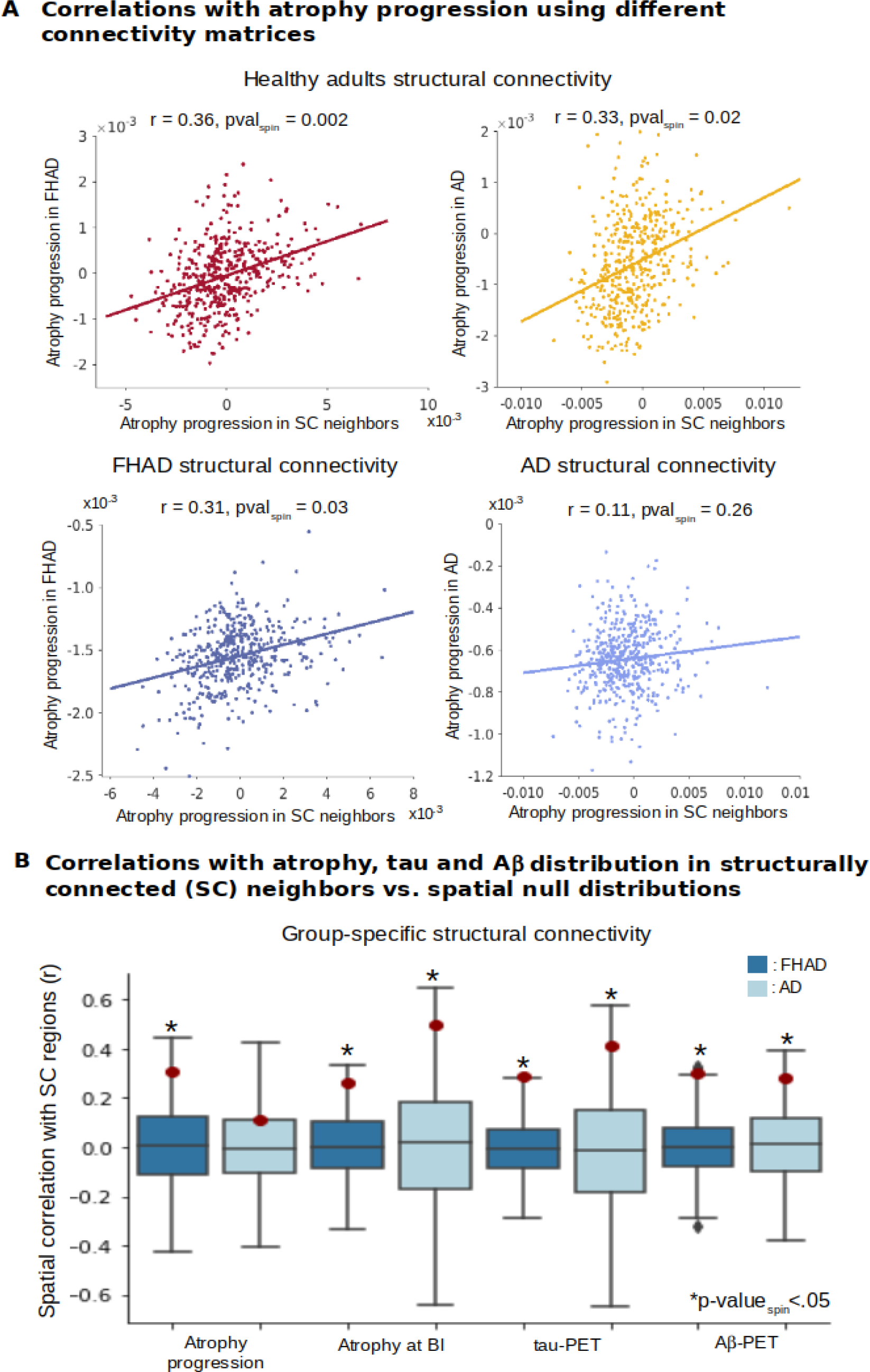
Relationships with structural connectivity in participants with a family history of Alzheimer’s disease (FHAD) and patients with Alzheimer’s disease (AD). (**A**) This panel shows the spatial correlations (Pearson’s r) between cortical atrophy progression in a given region and that in its structurally connected (SC) regions in both FHAD and AD. The upper row depicts correlations using a connectivity matrix from healthy adults, while the bottom row displays correlations using the group-specific connectivity matrices. (**B**) This panel illustrates the spatial correlations between brain atrophy progression, baseline atrophy, tau, and Aβ distribution in a region and those in its structurally connected regions, using group-specific structural connectivity matrices. In FHAD, all the correlations were significant while, in AD, only the correlation with atrophy progression was not significant. All correlations were tested against spatial null coefficient distributions.

#### Relationship with connectivity in FHAD

Using an healthy connectome, we found that the higher the baseline atrophy and atrophy progression, as well as the higher tau and Aβ concentration in a region, the higher the baseline atrophy (r=0.44, *p-value*_spin_=.001), atrophy progression (r=0.36, *p-value*_spin_=.02), tau (r=0.24, *p-value*_spin_=.009) and Aβ concentration (r=0.29, *p-value*_spin_=.003) in regions that are structurally connected (Supplementary Table.2). In contrast, baseline atrophy, atrophy progression, tau and Aβ concentration were lower in regions that were not structurally connected (baseline atrophy: r=-0.50, *p-value*_spin_=.001; atrophy progression: r=-0.36, *p-value*_spin_=.001; tau: r=-0.34, *p-value*_spin_=.001 and Aβ: r=-0.39, *p-value*_spin_=.001). Similar results were obtained using the structural connectivity matrix from FHAD (Fig.5A Left upper row; Supplementary Table.3). These results suggest that in FHAD, distributions of atrophy, tau, and Aβ all depend on structural connections in a similar manner.

#### Relationship with connectivity in AD

Using a structural connectivity matrix from healthy adults, a significant correlation was found between baseline atrophy in each region and that in their connected neighbors in AD (r=0.66, *p-value_spin_*=.001). Significant and positive correlations were also observed with atrophy progression (r=0.33, *p-value_spin_*=.02), tau (r=0.49, *p-value_spin_*=.001), and Aβ concentration (r=0.21, *p-value_spin_*=.004) (Fig.5A: Right upper row; Supplementary Table.2). Additionally, negative correlations were noted with the same measures when using the non-structurally connected neighbors (baseline atrophy: r=-0.67, *p-value*_spin_=.001; atrophy progression: r=-0.43, *p-value*_spin_=.001; tau: r=-0.36, *p-value*_spin_=.001 and Aβ: r=-0.32, *p-value*_spin_=.001). Using the structural connectivity matrix from AD, adjusted with ComBat for inter-site variability, a significant correlation was observed with baseline atrophy (r=0.50, *p-value_spin_*=.001). However, no significant correlations were found in AD between the atrophy progression in each region and that in their connected neighbors (r=0.11, *p-value_spin_*=.26) (Fig.5A: Right bottom row). Similarly, the correlation with atrophy progression in the non-structurally connected neighbors was also not significant (r=-0.16, *p-value_spin_*=.18). In addition, significant correlations were observed with tau (r=0.41, *p-value_spin_*=.03) and Aβ (r=0.28, *p-value_spin_*=.02) concentration in a region and the average concentration in their structurally connected neighbors (Fig.5B; Supplementary Table.3). These findings support that in AD, tau and Aβ accumulate longitudinally following the AD-specific structural connectome, whereas most of the atrophy progression rather depends on the healthy connectome, before AD-related damages have occurred.

#### Relationship with receptor and transporter distributions

Since regional vulnerability may influence the local vulnerability of regions to atrophy progression, we examined the spatial correlations between cortical atrophy progression in FHAD and AD and the distributions of seven different neurotransmitter receptors and transporters (Fig.6). A negative correlation between the distribution of the 5-HT6 receptors and atrophy progression in FHAD was observed (r=-0.15, *p-value_spin_*=.04), but it was not significant after FDR correction (r=-0.15, *p-value_spin-FDR_*=.19). A significant Pearson’s correlation against a spatial null model was found between the distribution of serotonin 5-HT6 receptors and atrophy progression in AD after FDR correction (r=-0.22, *p-value_spin-FDR_*=.04). No other significant correlations were observed with atrophy progression in both groups (Supplementary Table.4). The observed negative correlation between atrophy progression and serotonin 5-HT6 receptors in AD possibly reflects the ceiling effect and the lower atrophy progression at older age in AD.

**Figure 6.**
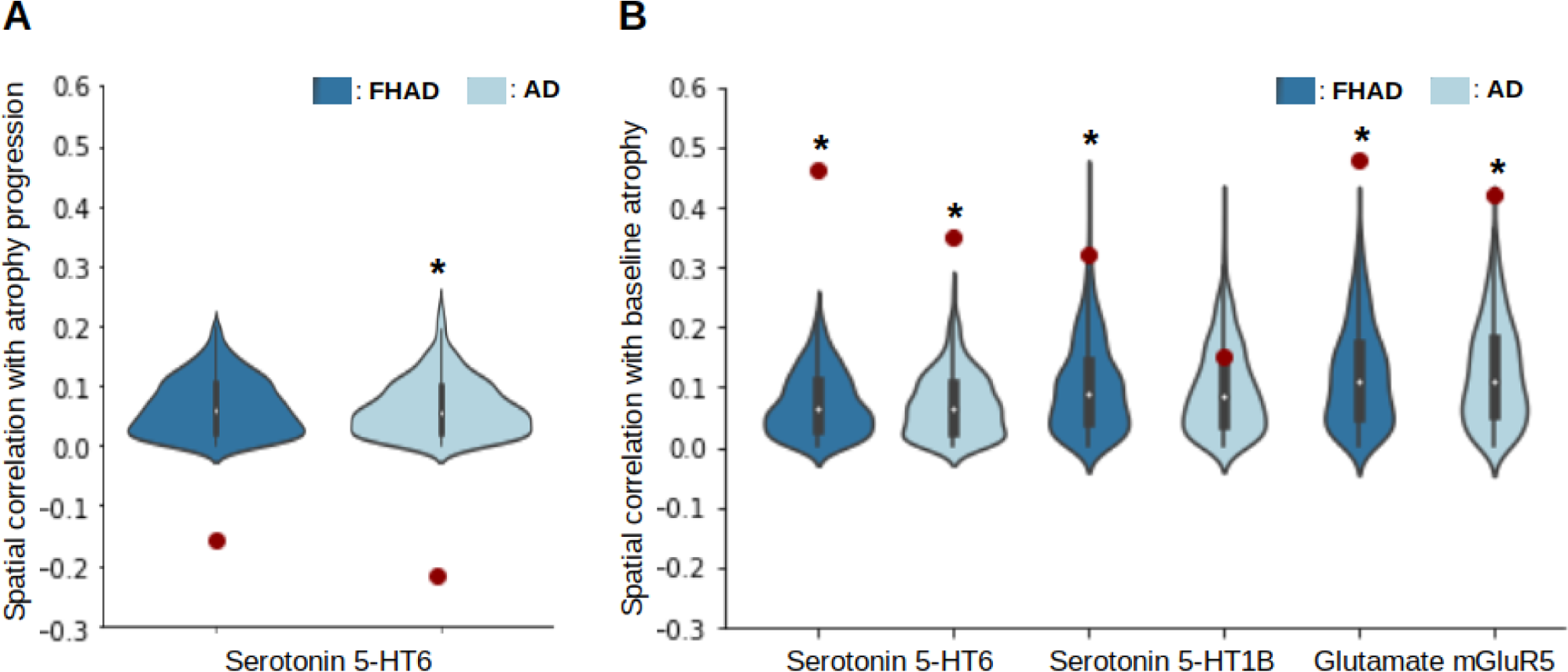
Serotonin and glutamate receptor distribution related to brain atrophy in individuals with a family history of Alzheimer’s disease (FHAD) and Alzheimer’s disease (AD). (**A**) The spatial distribution of the serotonin 5-HT6 receptor in cortical regions was negatively correlated with atrophy progression (β-values) in both FHAD and AD, but was only significant in AD after comparison with a spatial null distribution and FDR correction (\**p-value_spin-FDR_*<.05). (**B**) Positive and significant correlations were observed between the serotonin 5-HT6 receptor distribution and baseline atrophy in both groups, suggesting that the negative correlations most likely reflect a ceiling effect. In addition, the distribution of the serotonin 5-HT1B receptor was significantly correlated with baseline atrophy, but only in FHAD. Finally, significant correlations were observed between the glutamate mGluR5 receptor distribution and baseline atrophy in both groups.

As a supplementary analysis, the correlations with baseline atrophy in HC, FHAD, and AD, and the distributions of the neurotransmitter receptors and transporters were also investigated. Subsequent analysis showed positive and significant correlations between 5-HT6 distributions and atrophy at baseline in HC (r=0.45, *p-value_spin-FDR_*=.004), FHAD (r=0.46, *p-value_spin-FDR_*=.004), and AD (r=0.35, *p-value_spin-FDR_*=.004). This suggests that regions with more atrophy at baseline also possess higher concentrations of serotonin 5-HT6 receptors across all groups. A significant correlation was also observed with the distribution of serotonin 5-HT1B receptors in HC (r=0.33, *p-value_spin-FDR_*=.01) and FHAD (r=0.32, *p-value_spin-FDR_*=.01), but not in AD (r=0.15, *p-value_spin-FDR_*=.17). Additional analysis showed significant correlations between atrophy at baseline and the distribution of glutamate (mGluR5) receptors in HC (r=0.44, *p-value_spin-FDR_*=.004), FHAD (r=0.48, *p-value_spin-FDR_*=.004), and AD (r=0.42, *p-value_spin-FDR_*=.004). No other significant correlation was observed with atrophy at baseline (Supplementary Table.5). Taken together, these results suggest that more atrophy is observed in regions with a higher concentration of serotonin 5-HT6 and mGluR5 receptors in all groups, while a similar association with the serotonin 5-HT1B distribution is only observed in HC and FHAD.

## Discussion

We found similar patterns of cortical atrophy progression in FHAD and AD, although the severity and spatial extent were greater in AD. Tau and Aβ proteins also showed a similar regional distribution in both groups, but the concentration was higher in AD. Interestingly, the distribution of these proteins correlated spatially with atrophy at baseline, but not with the subsequent atrophy progression, possibly due to a lower atrophy progression with age (possible ceiling effect of atrophy). Indeed, it is likely that there is an upper limit on the amount of tissue loss detectable with MRI and brain deformation measure (DBM). Structural connectivity analyses revealed that both proteins accumulate in connected regions in both FHAD and AD. Brain atrophy patterns and their progression also aligned with existing structural connectivity in FHAD. However, in AD, most of the progression of atrophy rather depends on the healthy connectome, prior to the onset of AD-related white matter damage. In addition, brain regions with more atrophy at baseline were associated with higher levels of serotonin 5-HT6 and glutamate (mGluR5) receptors distributions derived from healthy older adults (HC), FHAD, and AD, while an association with serotonin 5-HT1B receptor distributions was only observed in HC and FHAD.

### Brain atrophy progression pattern in FHAD and AD

Our study revealed a similar cortical atrophy progression pattern in FHAD and AD, although with distinct differences in severity and regional distribution. Notably, a significant difference was observed in atrophy progression between the groups in 92 out of 448 cortical regions. Both groups exhibited significant atrophy progression in specific regions of the cingulate cortex, as well as the temporal and parietal cortices. This is consistent with studies reporting atrophy in the cingulate cortex in individuals with MCI and AD (Choo et al., 2010; Frisoni et al., 2002; Jones et al., 2006). These findings also are in alignment with prior research documenting accelerated atrophy rates in the temporal and parietal cortex in AD (Bakkour et al., 2013; Binette et al., 2020; Du et al., 2001; Fox et al., 2000; Jagust et al., 2008) and FHAD (Markus Donix et al., 2010; Kate et al., 2016). In addition, our data suggest a ceiling effect of the atrophy progression notably in AD, as detected by MRI, particularly in the precuneus and regions within the temporal, frontal, and occipital cortices. These findings align with previous studies, which report that atrophy often begins in the parietal and temporo-occipital cortex, in addition to frontal association areas, before spreading to other cortical regions (Apostolova & Thompson, 2008; Dickerson et al., 2011; Morris et al., 2009). Consistent with a ceiling effect, in AD, regions exhibiting higher baseline atrophy also showed less atrophy progression over time, a correlation that was not statistically significant in FHAD. This discrepancy could be attributed to the fewer regions (n=12) in FHAD showing negative atrophy progression compared to AD (n=102). Furthermore, negative correlations between the rate of atrophy, as well as cognitive decline, and age in cortical areas have previously been observed in individuals with mild cognitive impairment (MCI) and AD, with older individuals exhibiting less progression in both atrophy and cognitive decline (Fiford et al., 2018; Holland et al., 2012; Hrast et al., 2023). The neurobiological mechanims underlying these negative relationships with age are still unclear and might be influenced by a ceiling effect of atrophy, notably in AD. Overal, our findings suggest that while the initial regions affected may be similar, the severity and spatial extent of disease progression differs between FHAD and AD.

### Relationship with tau and Aβ concentration

Relationships were observed between baseline atrophy and both tau and Aβ concentration in FHAD and AD. This is in accordance with previous studies suggesting a relationship between tau and Ab deposition and volume loss (Ekman et al., 2018; Malpetti et al., 2022). However, there was a negative relationship between atrophy progression and regional tau and Aβ concentration, particularly in AD, which possibly reflects a ceiling effect in the detection of ongoing tissue loss. While the correlations between the atrophy pattern and tau-PET were consistently higher than with Aβ-PET, the differences between the correlations were not significant. Further investigations will be needed to confirm the stronger relationship between cortical atrophy and tau deposition compared with Aβ deposition (Malpetti et al., 2022). The FHAD and AD groups also showed similar patterns in the spatial distribution of abnormal tau and Aβ proteins (r >.90), but the concentrations were consistently higher in AD. These results support other findings suggesting an important relationship between tau and Aβ proteins in FHAD and AD and the atrophy progression patterns (Ozlen et al., 2022; Vogel et al., 2020).

### Structural connectivity mediates atrophy and proteins propagation

The differences in brain atrophy progression in specific networks between FHAD and AD may underlie the more pronounced cognitive dysfunction observed in AD despite comparable age and educational backgrounds. AD preferentially targets distinct neuronal networks, notably the limbic and associative regions of the cortex (Braak & Braak, 1991). Atrophic changes in AD were found to be most pronounced in the limbic network, followed by the DMN (Grothe et al., 2016). Our study revealed that while FHAD did not show significant brain atrophy in these networks, in AD, atrophy appeared to plateau in the default mode and limbic networks and continued to progress in the somatomotor network. Moreover, in this study, AD patients with greater atrophy at baseline in either the DMN, frontoparietal, or dorsal attention network also exhibited more pronounced cognitive deficits, as indicated by lower MoCA scores. This aligns with previous findings of altered activation patterns in the DMN, frontoparietal, and somatomotor networks during cognitive tasks in AD (Li et al., 2015). A meta-analysis on resting-state connectivity has also corroborated the alterations within the DMN and limbic networks (Badhwar et al., 2017). Although altered structural patterns in the dorsal attention network have been reported in AD (Qian et al., 2014) and its early stages (Wu et al., 2023), the relationship between these alterations and the cognitive deficits in the disease remains to be further explored. FHAD-related atrophy has also been localized to the precuneus, a core component of the DMN (Kate et al., 2016). Yet, no significant correlation was found between brain atrophy in any of the seven Yeo networks, including the DMN, and MoCA scores in FHAD. This is possibly due to the milder cognitive impairments exhibited by these individuals. While no specific network seems to be significantly affected in FHAD yet, these findings are supporting the important role of the DMN in AD-related atrophy and cognitive dysfunctions.

To further evaluate the role of brain networks in the progression of AD pathology we tested whether the spatial patterns of atrophy were shaped by structural connectivity. In both FHAD and AD, the connectivity analysis revealed that structural connectivity significantly influences the baseline distribution of tau, Aβ and atrophy regardless of the connectivity matrix used (i.e., group-specific or intact). These results are consistent with a previous study in ADNI showing that using a AD-specific connectome compared to an intact connectome did not significantly impact the spread of atrophy predicted by a diffusion model (Powell et al., 2018). Our finding also aligns with the network propagation of tau and Aβ observed in AD as reported previously (Lee et al., 2022; Vogel et al., 2020). Interestingly, in AD alone, Aβ distribution was less influenced by structural connectivity compared to tau deposition and atrophy at baseline. This supports prior studies, including (Franzmeier et al., 2020) and (Vogel et al., 2020), which have demonstrated that tau plays a key role in AD neurodegeneration and spreads via both functional and structural connectivity. These observations are in line with the Braak staging scheme suggesting that tau pathology spreads along connections between regions (Braak & Braak, 1991). Tau deposition has been shown to closely correlate with patterns of atrophy and cognitive dysfunction in AD (Bejanin et al., 2017; Harrison et al., 2019; Ossenkoppele et al., 2016). In our study, we observed that while Aβ proteins exhibited similar relationships with structural connectivity in both FHAD and AD, tau distribution and baseline atrophy were more influenced by structural connectivity in AD. This discrepancy could be due to the broader spatial extent of atrophy and tau distribution in AD.

In FHAD, we found that the pattern of brain atrophy progression aligned with existing structural connectivity. In contrast, in AD, no significant correlation was observed between atrophy progression and current structural connectivity. However, when using the structural connectivity from healthy adults, a significant correlation with the structurally connected regions was observed in both FHAD and AD. This suggests that in AD, atrophy may continue to propagate along pre-existing structural pathways, despite current damage to connectivity or that progression of atrophy results from propagating pathology that occurred much earlier, on an intact connectome. This finding is also in line with the observed ceiling effect of atrophy in AD, where the ability to detect tissue loss with MRI appears to slow down or stop in many regions. As a result, the correlation between current structural connectivity and regional atrophy progression is weaker than with the atrophy at baseline in AD. In contrast, localized white matter damage in FHAD does not seem sufficient to disrupt the progression pattern of atrophy. Indeed, the correlation between atrophy progression in a given region and its structurally connected neighboring region remained consistent, whether we used the FHAD-specific connectivity matrix or the matrix derived from healthy adults. Nonetheless, our findings align with the network spread hypothesis initially proposed by (Braak & Braak, 1991), emphasizing the critical role of structural connectivity in the distribution of pathological markers and atrophy progression in both FHAD and AD.

### Relationship with serotonin and glutamate receptors distribution

The spatial distribution of brain atrophy at baseline was associated with higher regional levels of serotonin 5-HT6 and the metabotropic glutamate subtype 5 receptors (mGluR5) distribution in HC, FHAD, and AD. A negative correlation was also detected between atrophy progression and the spatial distribution of serotonin 5-HT6, most likely reflecting a ceiling effect of atrophy detection in AD. There was also a positive association between baseline atrophy and the distribution of serotonin 5-HT1B receptors that was unique to FHAD and controls. Both 5-HT6 and 5-HT1B receptors play significant roles in memory and learning (Childers & Robichaud, 2005). Indeed, when inhibited, 5-HT6 receptors have been shown to enhance cognitive function (Mitchell & Neumaier, 2005; Perez-Garcia & Meneses, 2008). Moreover, antagonists of 5-HT1B receptors have been shown to improve memory and cognitive performance in high-cognitive-demand conditions (Buhot et al., 2000). Our findings are in line with these observations, revealing more atrophy in regions with higher 5-HT6 receptor densities in normal aging, FHAD, and AD, all conditions where different levels of memory decline occur (Koen & Yonelinas, 2014). The relationships found with 5-HT1B receptor distribution in normal aging and FHAD suggest that these receptors might contribute most significantly to atrophy in these conditions. In addition, elevated glutamate levels have been implicated in neurotoxicity and AD neurodegeneration (Cheng et al., 2021; Findley et al., 2019). Our study identified a positive correlation between atrophy distribution and higher regional levels of glutamate mGluR5 receptors in normal aging, FHAD, and AD. The mGluR5 receptors might contribute to Aβ toxicity through various mechanisms, including facilitating Aβ clustering (Renner et al., 2010). Furthermore, mGluR5 receptors may serve as a bridge between Aβ and tau pathology in AD, eventually leading to tau phosphorylation (Larson et al., 2012). The relationships with the mGluR5 receptor distribution found in this study supports the important role of this receptor in neurodegeneration not only in AD, but also in normal aging and FHAD.

### Strengths and limitations

This study has multiple strengths. It includes three demographically similar groups relative to age, sex and education, including a control group. This allows for a direct comparison between FHAD and AD in identifying both shared and distinct patterns of atrophy progression and underlying pathological mechanisms. This study also utilized a multi-modal approach (T1-weighted MRI, diffusion MRI, tau-PET, and Aβ-PET scans) to cover multiple facets of AD and FHAD, including atrophy patterns, structural connectivity effects, and protein deposition. This provides a holistic view of the brain atrophy progression in both FHAD and AD. A negative relationship between atrophy progression and age, possibly reflecting a ceiling effect in AD, was also identified, which could be crucial for understanding the extent to which atrophy can progress or be detected, and for adequately interpreting findings. In addition, the study extends beyond the commonly studied tau and Aβ proteins by exploring the role of different receptors and transporters in atrophy. This exploration encompasses not only AD but also FHAD and normal aging, thereby offering new insights for research in early intervention strategies.

A few limitations should also be noted. This study does not account for all potential confounders, such as medication use or lifestyle factors like physical, cognitive, and social activities, which might influence pathology progression. To mitigate the effect of potential confounding factors, statistical analyses were performed, controlling for age, education, BMI, sex and *APOe4* status. In addition, the study relies on one measure, DBM, for measuring atrophy, and DWI for structural connectivity. The inclusion of cortical thickness and surface area measurements, as well as functional MRI connectivity, could have enriched the findings. Another limitation is the study’s focus on cortical regions, which overlooks the role of subcortical areas in AD pathology. However, cortical atrophy patterns have been suggested to be more specific for monitoring preclinical and early stages of AD (Pini et al., 2016). Future studies could build upon our findings by incorporating multi-modal imaging and extending the analysis to subcortical regions to provide a more holistic understanding of the complex interplay between structural connectivity, protein deposition, and atrophy in FHAD and AD.

## Conclusion

This study delves into the pathological mechanisms at play in FHAD, highlighting both unique and shared neurodegenerative mechanism between FHAD and AD. While structural connectivity influences both atrophy and protein propagation in FHAD and AD, the extent of pathology appears to be less severe and less widespread in FHAD. This study also revealed that regions with higher levels of serotonergic and glutamatergic receptors may be especially susceptible to degeneration across normal aging, FHAD, and AD. These findings not only shed light on potential biochemical pathways contributing to neurodegeneration but also suggest that AD atrophy may be influenced by mechanisms present in both normal aging and FHAD, in addition to AD-specific pathology.

## Acknowledgments

We would like to thank Paul Cuciureanu for his help with the DWI data processing. We also thank Bratislav Misic and his team for making their maps of neurotransmitter receptors and transporters in addition to their connectivity matrices from healthy adults openly available.

## Supplementary material

Supplementary material is available online.

