## Supplementary Table. Supplementary Fig. for "Pattern and mechanisms of atrophy progression in individuals with a family history of Alzheimer’s disease: a comparative study"

***Supplemental Information***

**Figure 1.** Regions with significantly more (A) and less (B) atrophy progression between the participants with Alzheimer’s disease (AD) and individuals with a family history of Alzheimer’s disease (FHAD)

**A	Regions with significant atrophy progression**

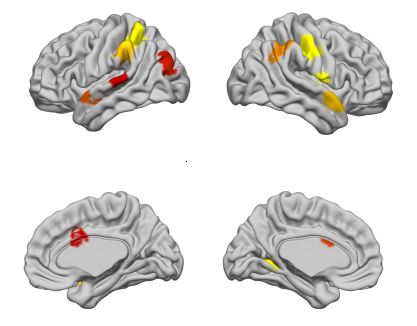

**B	Regions with less atrophy progression: possible ceiling effect**

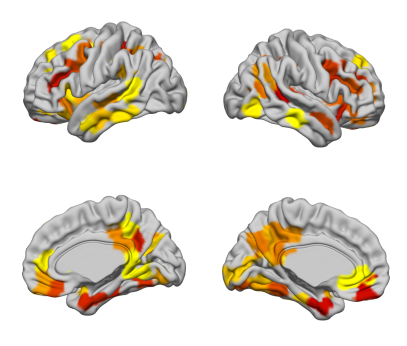

**Figure 2.** Beta values associated with the group effect in cortical regions showing significant higher tau (A) and beta-amyloide (Aβ) accumulation (B) in participants with Alzheimer’s disease (AD) vs. individuals with a family history of Alzheimer’s disease (FHAD)

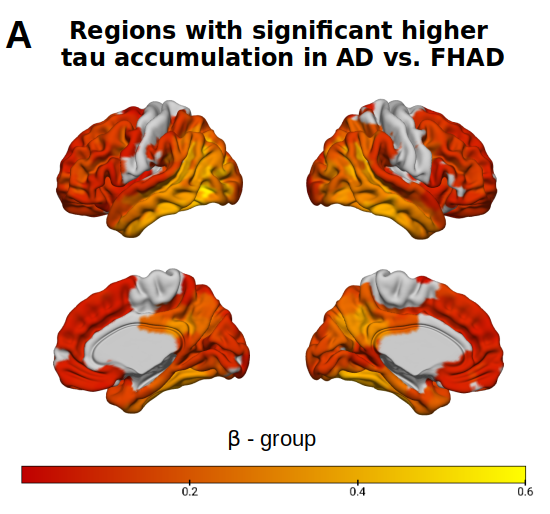

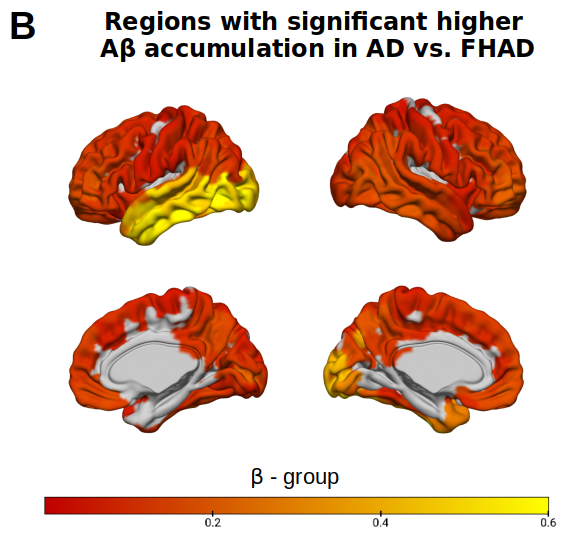

**Figure 3.** Regional Pearson’s correlations between atrophy at baseline, and tau (A) and Aβ accumulation (B) in participants with Alzheimer’s disease (AD) and individuals with a family history of Alzheimer’s disease (FHAD) using only participants who had both PET and MRI data at baseline

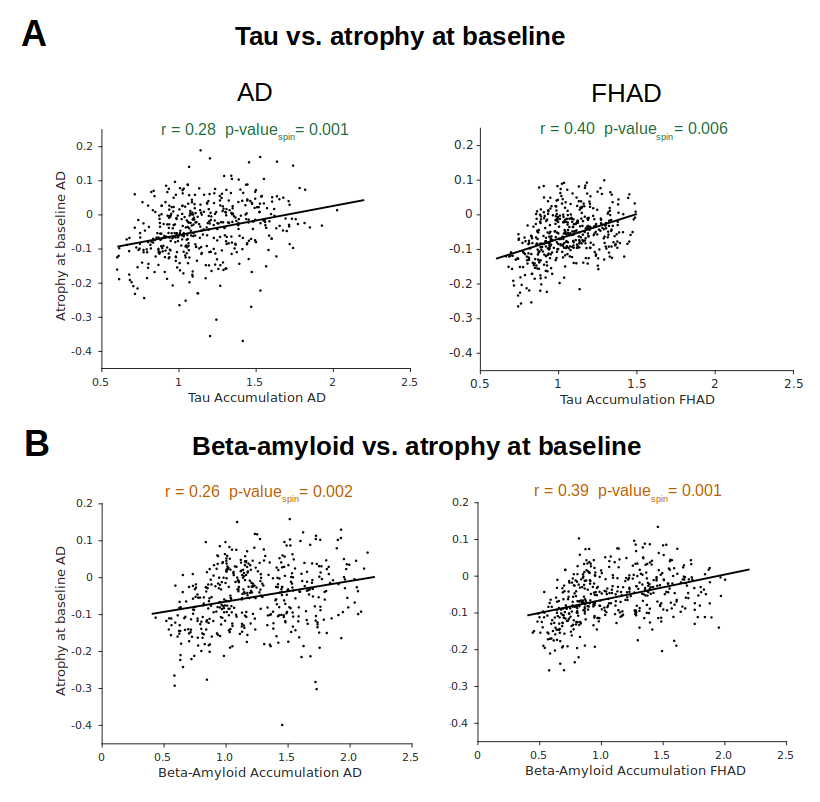

Table.1

**Number of subjects at each main step of the method for the participants with Alzheimer’s disease (AD), individuals with a family history of AD (FHAD) and hea**
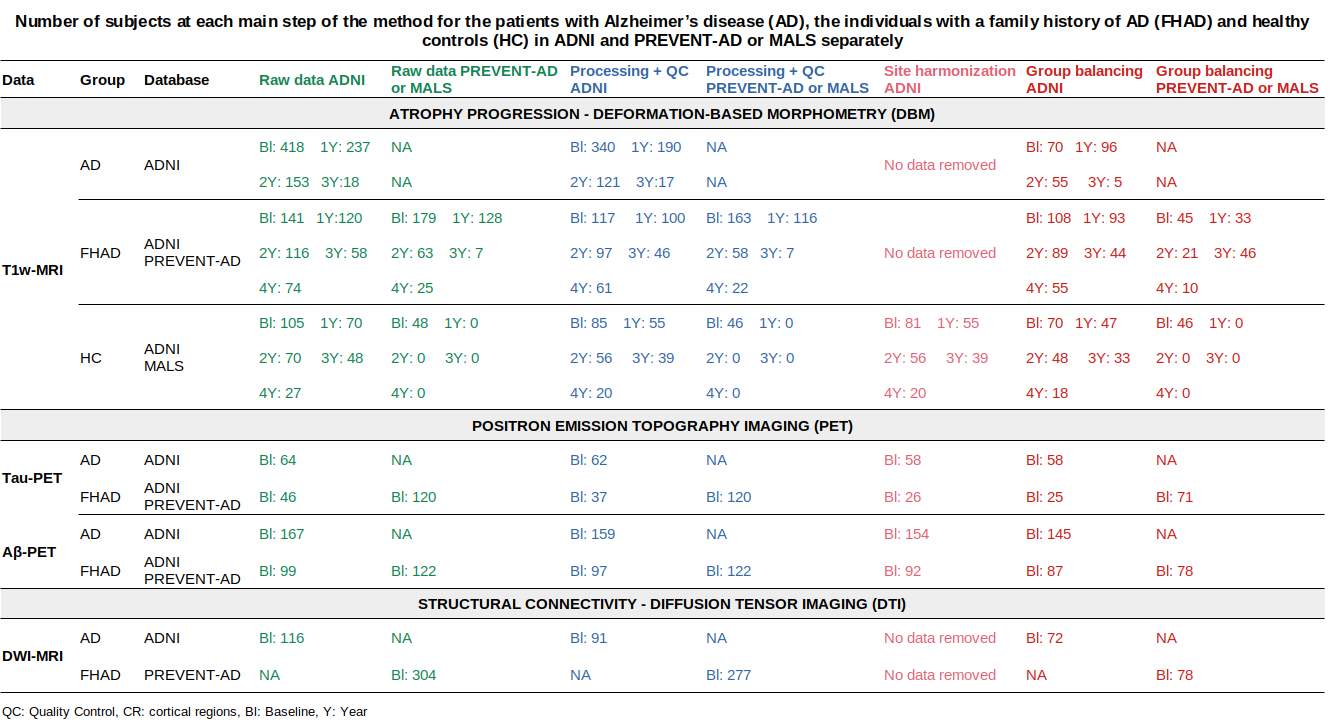
**lthy controls (HC) in the ADNI and PREVENT-AD or MALS database separately**

Table.2

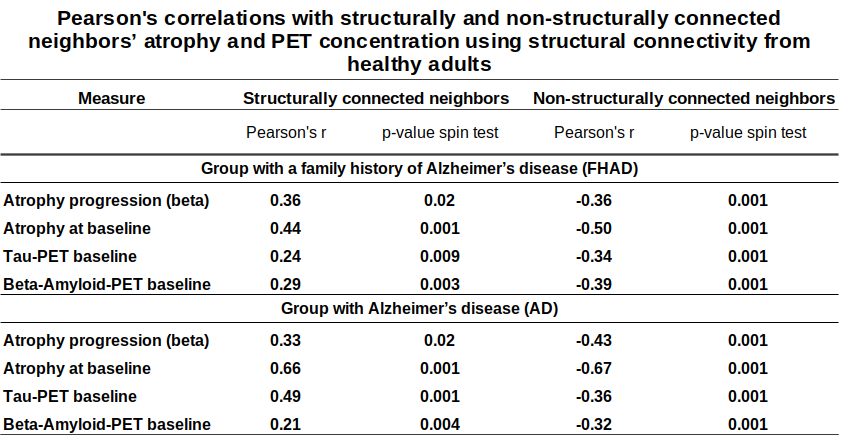

Table.3

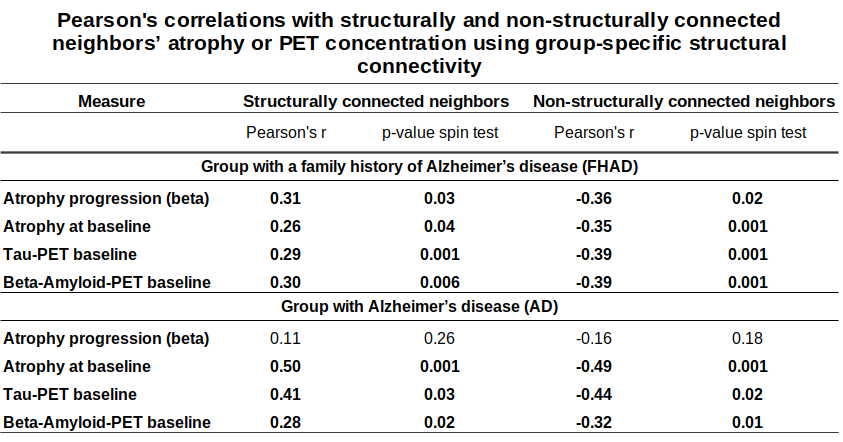

Table.4

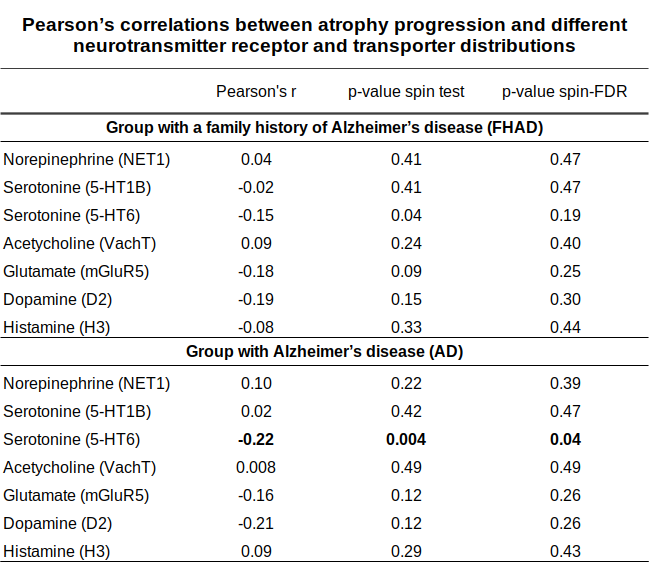

Table.5

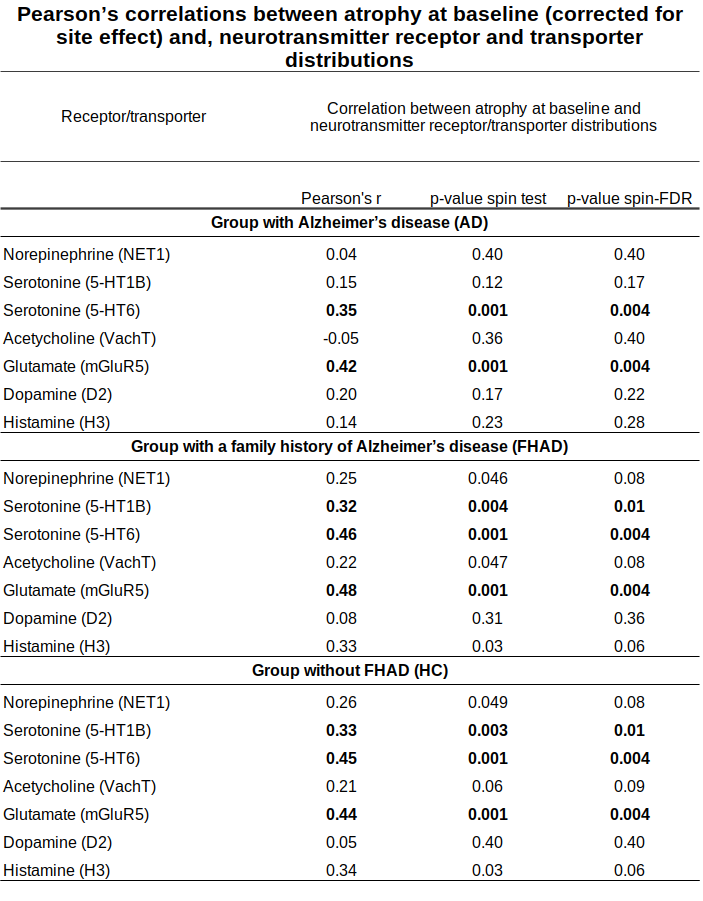
